# Treatment-seeking for children with suspected severe malaria attending community health workers and primary health centres in Adamawa State, Nigeria

**DOI:** 10.1101/2021.12.01.21267130

**Authors:** Tristan T. Lee, Elizabeth Omoluabi, Kazeem Ayodeji, Ocheche Yusuf, Charles Okon, Nina C. Brunner, Giulia Delvento, Aita Signorell, Mark Lambiris, Marek Kwiatkowski, Christian Burri, Christian Lengeler, Emmanuel Emedo, Fatima Cheshi, Manuel W. Hetzel

## Abstract

**Background:** The Community Access to Rectal Artesunate for Malaria project investigated the feasibility of introducing pre-referral rectal artesunate into existing community-based health services. In that study, the case fatality rate of children visiting primary health centres (PHCs) was 19% compared to 6% in children first visiting community health workers, locally called Community Oriented Resource Persons (CORPs). As case management practices did not fully explain this finding, this publication investigates other reasons underlying the observed difference in case fatality.

**Methods:** The observational study enrolled 589 children under the age of five years with fever and danger signs indicative of severe malaria attending CORPs and PHCs in Adamawa State, Nigeria, between June 2018 and July 2020. After 28 days, follow-up visits were conducted with caregivers to understand background characteristics, severity of symptoms, home treatment administration, and treatment seeking practices during the child’s illness. These factors were compared between children visiting CORPs versus those visiting PHCs as their first health provider.

**Results:** Children visiting PHCs were more likely to display danger signs indicative of central nervous system involvement (90% vs. 74%, p < 0.01) and have four or more danger signs (50% vs. 39%, p = 0.02). The delay between illness onset and visiting the community-based provider did not differ between children attending a CORP and children attending a PHC. PHC attendances more often lived in urban areas (16% vs 4%, p=0.01) and travelled farther to their first health provider, which was usually a community-based provider. Although practicing home treatment was common, especially among children attending PHCs (42% vs 33%, p=0.04), almost none of the children were given an antimalarial. PHCs were visited for their professionalism and experience while CORPs were visited for their low cost and because caregivers personally knew and trusted the provider.

**Conclusions:** Our comparison of children with suspected severe malaria seeking care from two kinds of community-based health care providers in Nigeria suggest that illness severity may be the primary driver behind the observed difference in case fatality rate.

## Background

Uncomplicated malaria can quickly progress to severe malaria and death if left untreated (1-3). Therefore, early diagnosis and prompt, effective treatment are essential to further reduce malaria mortality (4). Treatment of uncomplicated malaria is generally provided by primary health centres (PHC) and, in several countries in sub-Saharan Africa and beyond, by community health workers implementing the integrated community case management (iCCM) strategy (5).

Health workers at that level can conduct a malaria rapid diagnostic test (mRDT) or sometimes perform microscopy diagnosis of malaria, and provide oral artemisinin-based combination therapy (ACT) to children with uncomplicated malaria. Children with severe malaria have to be referred to health centres or hospitals with inpatient facilities that can manage severe malaria comprehensively. These referral facilities are often more difficult to reach, particularly for communities in remote rural areas.

In 2018 the World Health Organization (WHO) granted pre-qualification to two products of rectal artesunate (RAS) that could be used as a pre-referral treatment for cases of suspected severe malaria in children diagnosed at community level (6). In a clinical trial, pre-referral RAS was shown to reduce death or permanent disability when administered to children under the age of six years with suspected severe malaria who delayed referral to a higher level health facility (7).

Nigeria accounts for 27% of the world’s malaria cases and 23% of malaria deaths (8). In addition to having the world’s highest burden of malaria, the country is also challenged with high levels of poverty, a large rural population with few transportation options, and a poorly functioning health system (9). This situation makes prompt and appropriate treatment seeking for severe malaria difficult to achieve in many places. Considering the potential of RAS to improve children’s health outcomes in such a context, pre-referral RAS was added to the treatment guidelines of local community-based providers, including community health workers implementing iCCM, known locally as Community Oriented Resource Persons (CORPs), and primary health centers (PHCs) implementing the integrated management of childhood illnesses (IMCI). Even though the national treatment guidelines had already included RAS as pre-referral treatment option before the WHO pre-qualification (10), RAS was not widely used and not at all provided by CORPs.

In an observational study, the Community Access to Rectal Artesunate for Malaria (CARAMAL) project investigated the health impact of introducing RAS in remote populations in the Democratic Republic of the Congo, Nigeria, and Uganda (11). In Nigeria, the study did not find a positive health effect of introducing pre-referral RAS in three Local Government Areas (LGA) of Adamawa State. The case fatality rate (CFR) among children with suspected severe malaria during pre and post-RAS implementation phases were 4.2% and 16.1%, respectively. Meanwhile, the study found that the CFR was higher in children first attending a PHC (18.5%) compared to those first attending a CORP (5.7%). These differences in CFR may be attributable to factors such as the health condition at the time of treatment seeking, care seeking practices, and treatments provided. Two critical aspects of the continuum of care for severe malaria cases, namely referral completion and post-referral treatment, have been described in other manuscripts (Signorell et al, manuscript in preparation, (12)). But neither seem to provide an explanation of the difference in CFR between the two groups of patients (11).

Understanding the reasons underlying these differences in CFR is crucial to ensure that appropriate measures are taken to reduce child deaths at each level of care. In this article, we therefore investigate whether there are underlying differences between children who visited CORPs and those who visited PHCs as community level providers. Currently, the literature lacks studies comparing the characteristics of children seeking treatment from different community-based providers. We specifically focused on the severity of symptoms, home treatment and treatment seeking delay, while taking into account background characteristics of the patients.

## Methods

### Study design

This research was conducted as part of the CARAMAL project that accompanied the roll-out of pre-referral RAS into existing iCCM and IMCI systems. The complete methods are detailed elsewhere (Lengeler et al., manuscript in preparation, (11)). This analysis includes children with suspected severe malaria enrolled by community-based providers (CORPs or PHCs) in Nigeria.

### Setting

Data was collected in Fufore, Mayo-Belwa and Song LGAs of Adamawa State in northeastern Nigeria. The estimated total population of these LGAs was 746,949 in 2018, of which 130,430 (17%) were under the age of five years (13). The population in the three LGAs is served by over 500 CORPs, 77 PHCs, and 3 referral health facilities (Cottage Hospitals). In the study area, malaria occurs seasonally.

CORPs and PHCs provide primary health care services at community level. Both use similar guidelines for the assessment and treatment of childhood illnesses, with CORPs implementing iCCM and PHCs implementing IMCI guidelines. PHCs are small health facilities with nurses and intermediate staff. These staff have an educational background in health, in contrast to CORPs who may not have a medical background. CORPs are provided with medical supplies and supervision through nearby PHCs. CORPs were introduced in 2014 to fill an accessibility gap and were placed in communities more than 5 km away from a health facility and in communities hard to reach due to bad road conditions, or natural barriers like rivers and mountains.

### Data collection

Children under five years visiting a PHC or a CORP with a history of fever and at least one danger sign indicative of suspected severe malaria according to Nigerian iCCM guidelines (unusually sleepy or unconscious, not able to drink or feed, vomiting everything, convulsions, or yellow eyes) were enrolled between June 2018 and July 2020. CARAMAL staff attempted to reach community-based providers weekly by phone calls or text message to identify and register children meeting the inclusion criteria. Follow-up visits were done 28 days later in person at the child’s residence, or by phone during the COVID-19 pandemic lockdown. Caregiver interviews were conducted in the local language (Hausa or Fulfulde) to collect information about the child’s health status, signs and symptoms of disease, treatment seeking perceptions and practices, and medicines the child received. Geolocations of the child’s house were recorded. Children who died were followed up two to three months after enrolment to respect the mourning period. Additional details about the circumstances of the death were asked. Data was collected electronically on tablets using ODK Collect, and saved on a password protected ODK Aggregate server hosted at the Swiss Tropical and Public Health Institute in Switzerland.

### Data analysis

Study participants were included in the analysis if they were successfully followed-up on day 28, their caregiver provided consent, and children were under the age of five, had a history of fever and a Nigeria specific iCCM danger sign, and had a positive mRDT test at enrolment. For consistency with complementary publications (11), children who died after more than 31 days were excluded from the analysis.

This study compares children attending a CORP with children attending a PHC. The main variables of interest were background characteristics, home treatment, treatment seeking delay, disease severity and reasons for going to the CORP or the PHC. Treatment seeking delay was defined as the reported number of days between illness onset and attending the CORP or PHC, categorized into two-day periods. We used the presence of danger signs involving the central nervous system (convulsions, unusually sleepy or unconscious), number of danger signs (out of convulsions, unusually sleepy or unconscious, vomits everything, unable to drink or feed, unable to sit or stand, blood in stool, swelling of both feet), and caregiver-perceived severity as proxies for disease severity. RAS implementation started in May 2019. COVID-19 movement restrictions were in place between April 1, 2020 and the end of the study. The rainy season usually lasts from May to October, and was defined as such. Living in a rural or urban setting was determined based on the child’s residence geolocation and the LandScan HD: Nigeria version 1.1 geodatabase (Oak Ridge National Laboratory, 2018) per settlement patterns defined by Weber et al. (14).

Descriptive analyses comparing CORP and PHC attendance were done using logistic regression models with clustered standard errors (clustering at the level of the enrolling health care provider). These were adjusted for the effect of the COVID-19 pandemic as the pandemic was likely to affect treatment seeking. P-values were calculated using the likelihood ratio test for categorical variables and on the odds ratio for binary variables. P-values of <0.05 were considered statistically significant. Unadjusted models, sensitivity analysis using Generalized Estimating Equations, and a further explanation on statistical methods are presented in the supplementary materials. Statistical analyses were conducted in Stata SE version 16.1.

## Results

### Study population

CORPs and PHCs provisionally enrolled 724 children. Of those, 66 (9%) were lost to follow-up or the caregiver did not provide consent. We excluded another 69 (10%) children due to children not having a history of fever (n=7), no danger sign (n=26), dying > 31 days after enrolment (n=2), or no positive mRDT at enrolment (n=34) (Figure S1). Thus, the analysis included 589 children with suspected severe malaria, of which 314 (53%) were enrolled by CORPs and 275 (47%) by PHCs (Table 1).

**Table 1.** Patient characteristics by type of provider visited

|  | CORP |  | PHC |  | AOR* | 95% CI* | P-value* |
| --- | --- | --- | --- | --- | --- | --- | --- |
|  | N | % | N | % |  |  |  |
| **Total** | **314** |  | **275** |  |  |  |  |
| **Child sex** |  |  |  |  |  |  |  |
| Male | 188 | 60 | 164 | 60 | Ref. |  |  |
| Female | 126 | 40 | 111 | 40 | 1.0 | (0.7 - 1.3) | 0.91 |
| **Child age (years)** |  |  |  |  |  |  |  |
| 0 | 40 | 13 | 27 | 10 | Ref. |  | 0.34 |
| 1 | 86 | 27 | 74 | 27 | 1.3 | (0.7 - 2.6) |  |
| 2 | 82 | 26 | 82 | 30 | 1.5 | (0.8 - 2.9) |  |
| 3 | 68 | 22 | 55 | 20 | 1.3 | (0.6 - 2.4) |  |
| 4 | 38 | 12 | 37 | 13 | 1.5 | (0.7 - 3.4) |  |
| **Caregiver sex** |  |  |  |  |  |  |  |
| Male | 102 | 32 | 95 | 35 | Ref. |  |  |
| Female | 212 | 68 | 180 | 65 | 0.9 | (0.6 - 1.3) | 0.48 |
| **Caregiver age** |  |  |  |  |  |  |  |
| < 25 | 42 | 13 | 46 | 17 | Ref. |  | 0.17 |
| 25 - 34 | 167 | 53 | 128 | 47 | 0.7 | (0.4 - 1.2) |  |
| 35 - 45 | 75 | 24 | 72 | 26 | 0.8 | (0.5 - 1.5) |  |
| ≥ 45 | 30 | 10 | 28 | 10 | 0.9 | (0.4 - 1.7) |  |
| Missing | 0 | 0 | 1 | 0 | - |  |  |
| **Caregiver education\*\*** |  |  |  |  |  |  |  |
| Never attended school | 54 | 17 | 65 | 24 | Ref. |  | 0.77 |
| Primary or lower | 15 | 5 | 26 | 9 | 1.5 | (0.6 - 3.4) |  |
| Secondary or higher | 28 | 9 | 44 | 16 | 1.3 | (0.6 - 2.6) |  |
| Quranic | 22 | 7 | 39 | 14 | 1.5 | (0.7 - 3.1) |  |
| Missing | 195 | 62 | 101 | 37 | - |  |  |
| **LGA** |  |  |  |  |  |  |  |
| Fufore | 143 | 46 | 94 | 34 | Ref. |  | 0.02 |
| Mayo-Belwa | 95 | 30 | 154 | 56 | 2.3 | (0.5 - 10.4) |  |
| Song | 76 | 24 | 27 | 10 | 0.5 | (0.1 - 1.9) |  |
| **Residence** |  |  |  |  |  |  |  |
| Rural | 298 | 95 | 227 | 83 | Ref. |  |  |
| Urban | 14 | 4 | 43 | 16 | 4.6 | (1.4 - 15.4) | 0.01 |
| Missing | 2 | 1 | 5 | 2 | - |  |  |
| **Day of enrolment** |  |  |  |  |  |  |  |
| Workday | 241 | 77 | 239 | 87 | Ref. |  |  |
| Weekend | 73 | 23 | 36 | 13 | 0.5 | (0.3 - 0.9) | 0.02 |
| **Season** |  |  |  |  |  |  |  |
| Dry season | 83 | 26 | 66 | 24 | Ref. |  |  |
| Rainy season | 231 | 74 | 209 | 76 | 1.0 | (0.6 - 1.6) | 0.95 |
| **RAS implementation phase** |  |  |  |  |  |  |  |
| Pre-RAS | 156 | 50 | 61 | 22 | Ref. |  |  |
| Post-RAS | 158 | 50 | 214 | 78 | 3.3 | (1.7 - 6.4) | <0.01 |
| **COVID-19 pandemic** |  |  |  |  |  |  |  |
| Pre-COVID-19 | 268 | 85 | 207 | 75 | Ref. |  |  |
| COVID-19 period | 46 | 15 | 68 | 25 | 1.9 | (1.0 - 3.6) | 0.04 |

\*Logistic regression adjusting for the COVID-19 pandemic, with standard errors clustered at the level of the health care provider. Likelihood ratio test used to calculate p-values for categorical variables. \*\*Data not collected during the entire study period.

Children were enrolled from 139 community-based providers in the study area. The number of enrolled patients per provider varied from 1 to 42 (median=2, IQR: 2 – 4), with fewer PHCs and a tendency for larger numbers in PHCs. Heterogeneity of almost all variables was high within children enrolled by the same health provider.

### Patient characteristics

Overall, there were more male than female patients in the study and the mean patient age was 2.0 years. Caregivers were most often female and between the ages of 25 to 34. More children were enrolled in the rainy season than the dry season. There were no differences in the aforementioned aspects between patients enrolled by CORPs and PHCs (Table 1). Children first attending PHCs were more likely to live in an urban area (adjusted odds ratio [AOR]: 4.6, 95%CI: 1.4 – 15.4) and less likely to visit the facility during a weekend (AOR: 0.5, 95%CI: 0.3 – 0.9). They were also more often enrolled after the introduction of RAS in the study area (AOR: 3.3, 95%CI: 1.7 – 6.4), and during the COVID-19 pandemic (AOR: 1.9, 95%CI: 1.0 – 3.6).

### Signs and symptoms

Convulsions (79%) and being unusually sleepy or unconscious (70%) were more common in children visiting a PHC (Figure 1). Together, these two symptoms involving the Central Nervous System (CNS) were reported more frequently in children first visiting a PHC (90%) than in children first visiting a CORP (74%) (AOR: 3.5, 95%CI: 1.9 – 6.1). There was no significant difference in the proportion of children with CNS symptoms during the COVID-19 pandemic (79%) compared to prior to the pandemic (82%).

**Figure 1.**
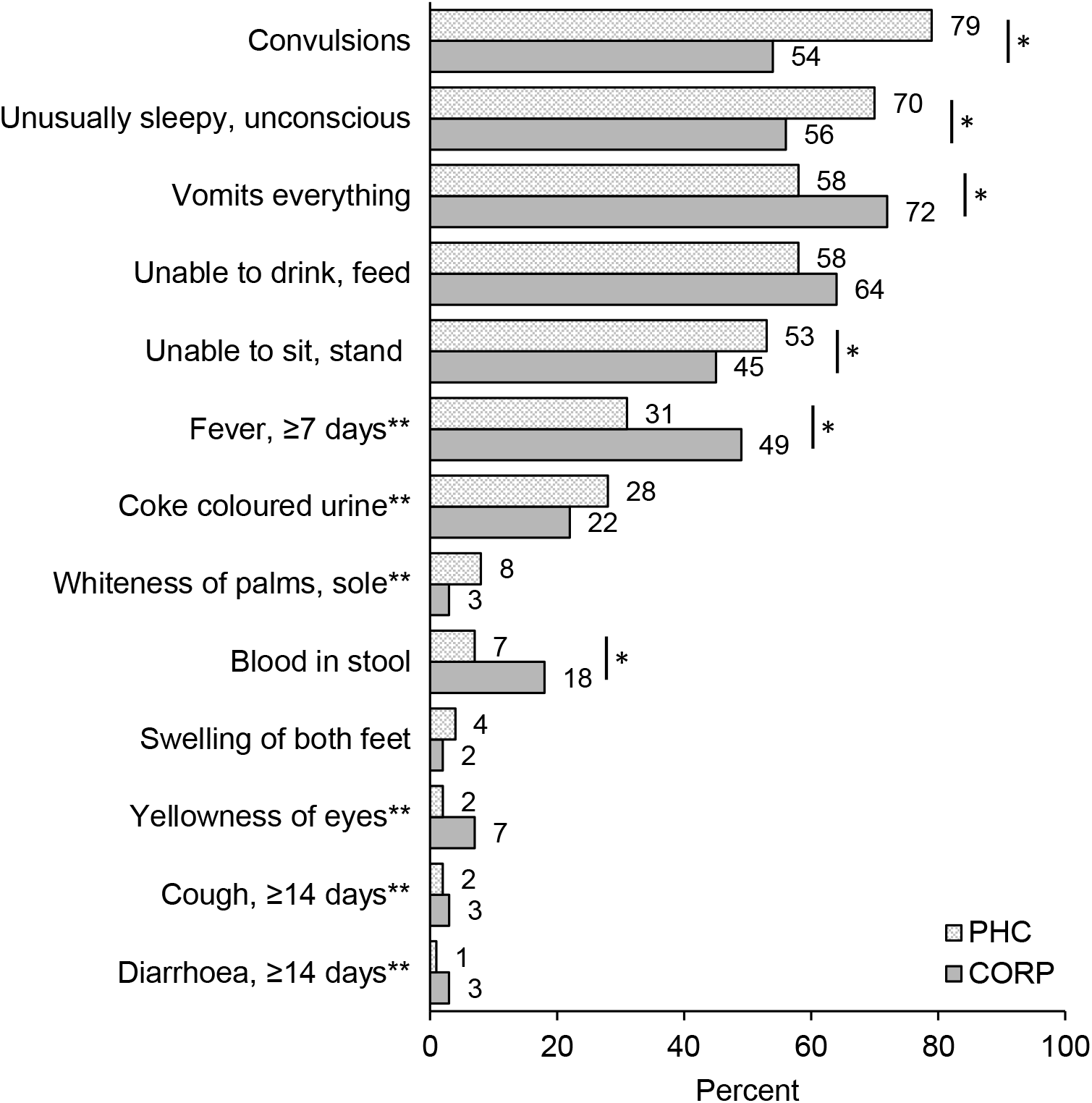
Signs and symptoms of severity during the child’s case of severe illness stratified by first community-based health provider visited. ^*^Considered statistically significant at p<0.05. ^**^Data not collected during the entire study period.

Children enrolled at CORPs were more often reported to vomit everything (72%) or be unable to drink or feed anything (64%). Vomiting everything, prolonged fever (≥7 days) and blood in the stool were reported significantly more frequently among children first attending a CORP (Figure 1).

Children often presented with multiple symptoms: proportionately more children attending a PHC (50%) had ≥ 4 danger signs compared to children taken to a CORP (39%) (p=0.02) (Table 2). Danger signs that were often reported together included convulsions and being unusually sleepy/unconscious (CORP: 36%, PHC: 59%), being unusually sleepy/unconscious and not being able to eat/drink (CORP: 39%, PHC: 46%), being unable to eat/drink and being unable to sit/stand (CORP: 35%, PHC: 41%). The following combination of three danger signs was also commonly reported: convulsions, unusually sleepy/unconscious, and unable to eat/drink anything (32% overall, 26% among CORPs visits, 39% among PHC visits).

**Table 2.** Number of danger signs and caregiver’s perceived severity of the illness

|  | CORP |  | PHC |  | AOR* | 95% CI* | p-value* |
| --- | --- | --- | --- | --- | --- | --- | --- |
|  | N | % | N | % |  |  |  |
| **Total** | **314** |  | **275** |  |  |  |  |
| **Number of danger signs \*\*\*** | **314** |  | **275** |  |  |  |  |
| 0 - 1 | 52 | 17 | 32 | 12 | Ref. |  | 0.02 |
| 2 - 3 | 138 | 44 | 106 | 39 | 1.4 | (0.8 - 2.4) |  |
| ≥ 4 | 124 | 39 | 137 | 50 | 2.0 | (1.2 - 3.4) |  |
| **Perceived severity of the illness** |  |  |  |  |  |  |  |
| Not fatal | 236 | 75 | 193 | 70 | Ref. |  |  |
| Fatal | 76 | 24 | 79 | 29 | 1.3 | (0.8 - 2.0) | 0.25 |
| Don't know/Missing | 2 | 1 | 3 | 1 | - |  |  |

\*Logistic regression adjusting for the COVID-19 pandemic, with standard errors clustered at the level of the health care provider. Likelihood ratio test used to calculate p-values for categorical variables. \*\*3 children were not said to have any danger signs by caregivers at follow-up but health providers noted danger signs at enrolment. \*\*\*Only includes danger signs collected across the whole study period.

In three-quarters of the cases, caregivers perceived their child’s illness to be serious but not fatal with no statistically significant difference between the two compared groups.

### Home treatment

Home treatment was more common among patients attending a PHC than those attending a CORP (AOR: 1.5, 95%CI: 1.0 – 2.1) (Table 3). Treatments included modern medicines, traditional medicines or herbs, and washing or sponging to cool the fever. Caregivers who were able to name the medicine most often gave paracetamol (CORP: 85%, PHC: 92%) and occasionally artemether-lumefantrine, the first-line antimalarial drug (CORP: 15%, PHC: 14%). The type of home treatment did not differ between the two groups.

**Table 3.** Actions taken at home prior to consulting health providers

|  | CORP |  | PHC |  | AOR* | 95% CI* | P-value* |
| --- | --- | --- | --- | --- | --- | --- | --- |
|  | N | % | N | % |  |  |  |
| **Total** | **314** |  | **275** |  |  |  |  |
| **Any home treatment** |  |  |  |  |  |  |  |
| No | 209 | 67 | 157 | 57 | Ref. |  |  |
| Yes | 103 | 33 | 116 | 42 | 1.5 | (1.0 - 2.1) | 0.04 |
| Missing | 2 | 1 | 2 | 1 | - |  |  |
| **Actions taken** | **103** |  | **116** |  |  |  |  |
| Traditional medicines/herbs | 22 | 21 | 26 | 22 | 1.1 | (0.5 - 2.2) | 0.86 |
| Tepid sponging | 7 | 7 | 17 | 15 | 2.4 | (0.9 - 6.4) | 0.09 |
| Given medicine | 79 | 77 | 85 | 73 | 0.8 | (0.5 - 1.5) | 0.55 |
| **Medicines given (if known)** | **67** |  | **76** |  |  |  |  |
| Artemether-lumefantrine | 10 | 15 | 11 | 14 | 1.0 | (0.3 - 3.2) | 0.99 |
| Oral rehydration solution (ORS) | 1 | 1 | 1 | 1 | 0.8 | (0.1 - 13.4) | 0.89 |
| Paracetamol | 57 | 85 | 70 | 92 | 2.1 | (0.6 - 7.0) | 0.23 |
| Other | 6 | 9 | 5 | 7 | 0.7 | (0.2 - 2.5) | 0.56 |

\*Logistic regression adjusting for the COVID-19 pandemic, with standard errors clustered at the level of the health care provider.

### Treatment seeking

Further details on treatment seeking were collected for 264 children who attended a CORP and 242 who attended a PHC. Of these, 14% of children attending a PHC and 7% of those attending a CORP had previously been to another provider (AOR: 2.2, 95%CI: 1.1 – 4.4). In most cases, the previous provider was a chemist/pharmacy/drug shop for both CORP (47%, 9/19) and PHC (71%, 24/34) enrolments (Fisher’s test p=0.85).

There was no substantial difference overall in treatment seeking delay to the community-based provider between children first visiting CORPs compared to those first visiting PHCs (Figure 2A). Across the study period, in children seeing CORPs, 38% of children sought care the same day or the day after becoming ill and 32% sought care 2-3 days after becoming ill. In PHC enrolments, slightly more children went to the enrolling provider 2-3 days after symptom onset (41%) compared to the same day or day after (35%). Home treatment was also associated with delay to a community-based provider. Children seeking care promptly had the lowest rates of home treatment administration (CORP: 18% and PHC: 32%) compared to those with greater delays (CORP: 44% and PHC: 52% in delays greater than one day) (p<0.01). The COVID-19 pandemic was found to decrease timely treatment seeking (same or following day) to CORPs (AOR: 5.1, 95%CI: 1.8 – 14.3) but not to PHCs (AOR: 1.5, 95%CI: 0.7 – 3.1).

**Figure 2.**
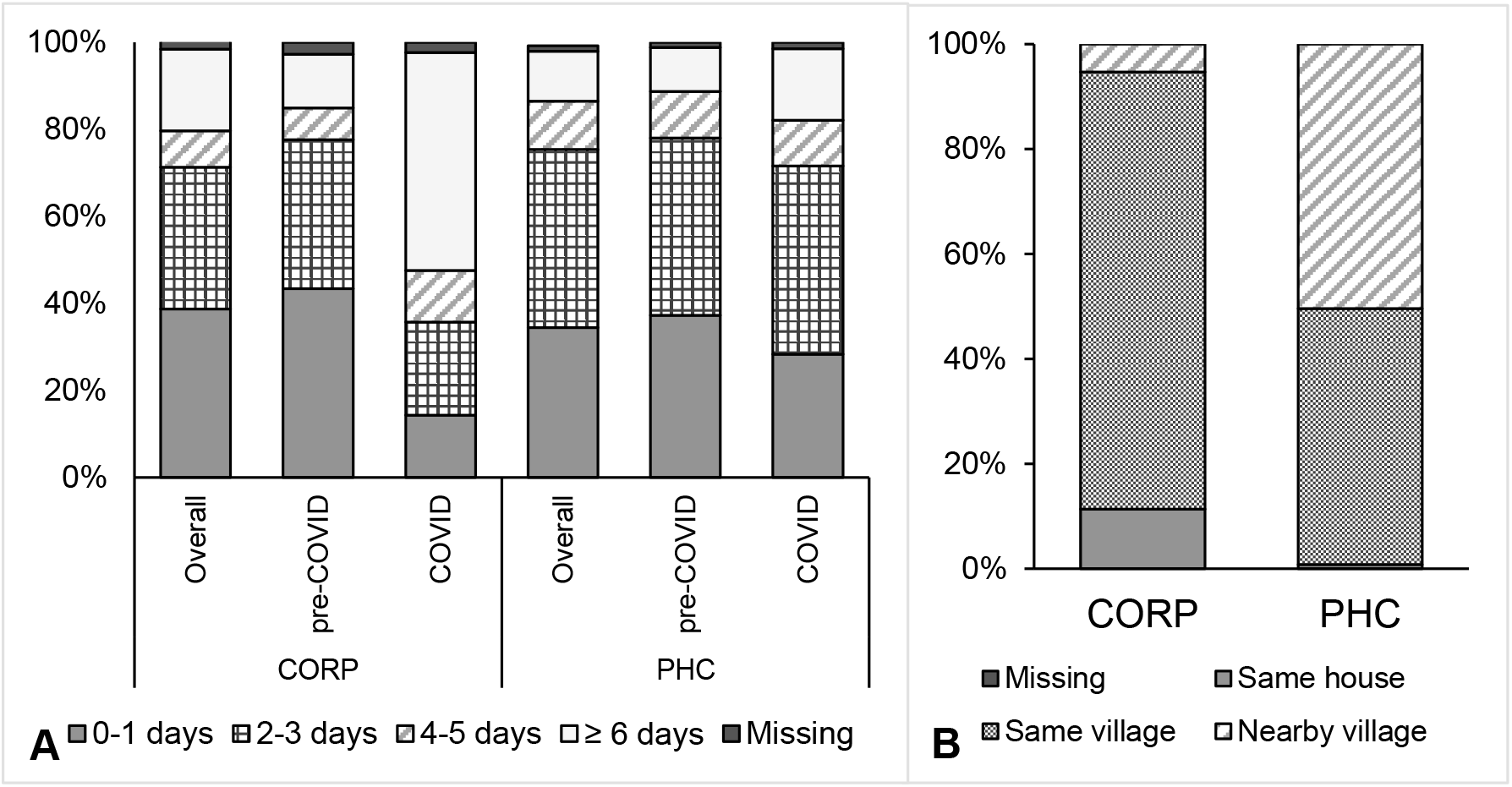
Treatment seeking to the first community-based provider: A) Treatment seeking delay between the time of illness onset and going to the enrolling provider in days by enrolment period. B) Location of the enrolling provider in relation to the child’s residence.

PHCs, offering a wider range of services and being much fewer than CORPs, were significantly more likely to be located outside the child’s village than CORPs (50% vs. 6%; AOR: 18.0, 95%CI: 6.8 – 47.2) (Figure 2B).

A total of 456 caregivers provided reasons for taking their child to the CORP or the PHC (Figure 3). The most common reasons for visiting a CORP were knowing (76%) and trusting (26%) the provider and low cost (22%). In contrast, reasons for attending a PHC included the experience (49%) and medical professionalism (34%) of PHC health workers, as well as knowing the provider (32%). Caregivers who brought their child to a CORP were significantly more likely to list knowing or trusting the provider, and the service being inexpensive as reasons for visiting, whereas those attending a PHC were more likely to list the experience and professionalism of the provider, and an expectation or custom to consult the provider as reasons.

**Figure 3.**
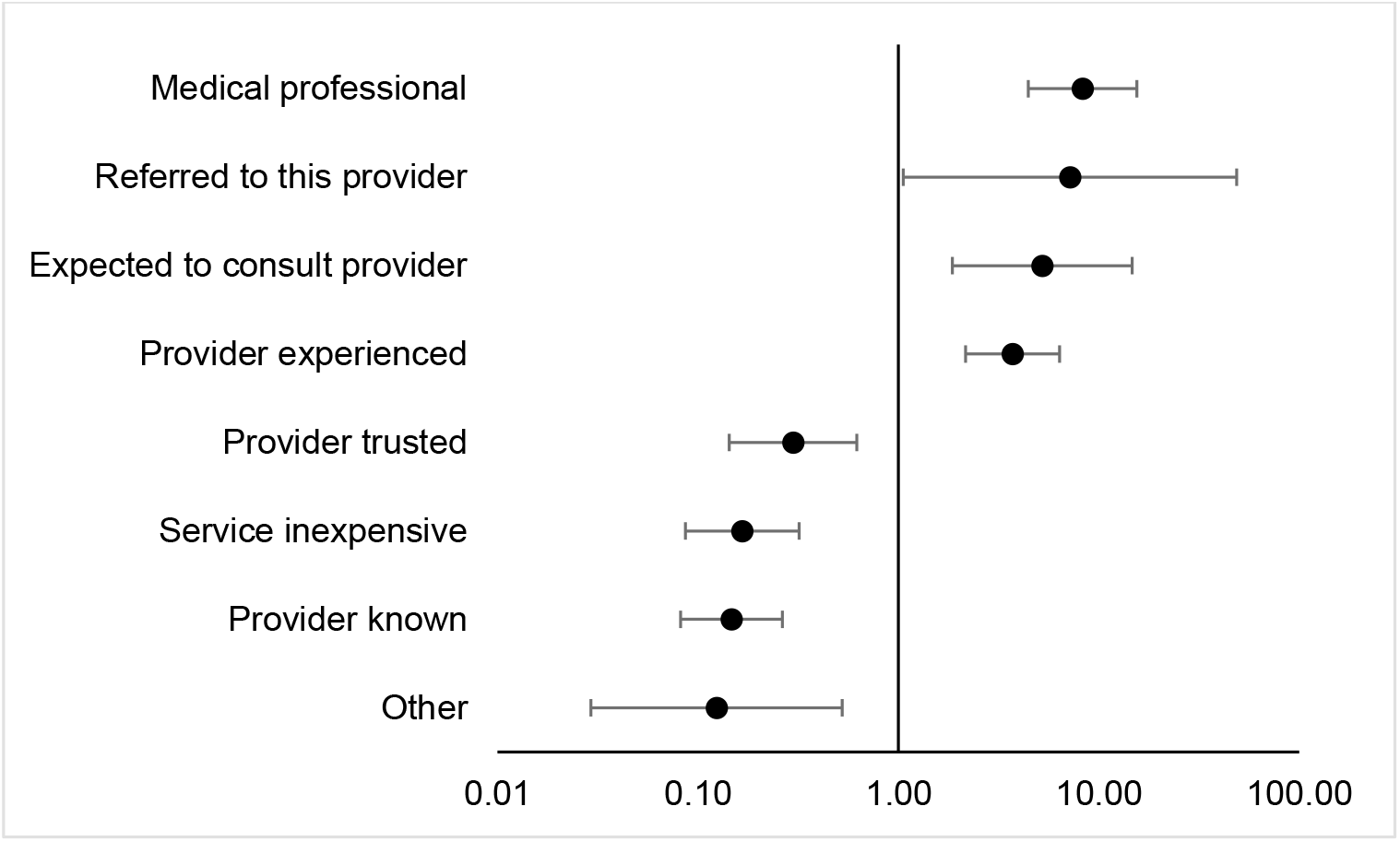
Odds ratio for reasons caregivers took their child with severe febrile illness to the initial CORP compared to the PHC provider. Logistic regression adjusting for the COVID-19 pandemic, with standard errors clustered at the level of the health care provider.

## Discussion

Early diagnosis and prompt effective treatment are critical in the management of malaria to prevent progression to severe disease, lasting disability, and death (4). We previously found that children with suspected severe malaria who first attended a PHC were three times more likely to die than those who visited a CORP (CFR 18.5% vs. 5.7%), a finding that could not be explained by differences in referral delay or post-referral treatment (11). Here, we investigated differences in the characteristics and treatment history of patients relative to their visit to the first community provider (CORP or PHC).

There were no differences in the demographic characteristics of patients attending CORPs or PHCs. However, children who first saw PHCs appeared to be more severely ill than those who first saw CORPs as reflected in the presence of CNS danger signs (90% vs. 74%) and the number of reported danger signs. The lack of an expert clinical examination of all patients included in this study limited the extent to which differences in illness severity could be examined, although using presence of CNS symptoms as a proxy for severe disease has been done previously (15-18). Additionally, caregivers were asked about signs and symptoms at any point during their child’s complete illness course. Thus, the causal relationship between severity and the complexity of disease progression and treatment seeking is difficult to elucidate. Warsame *et al*. (2016) previously found that children attending community health workers, in settings comparable to ours, had higher rates of CNS symptoms compared to those attending other health providers such as trained mothers or traditional healers, suggesting that children with more severe symptoms may be more likely to be taken to particular health providers, possibly based on the perceived professional capacity of the providers (18). In Burkina Faso, cases of severe disease often bypass community health workers in favour of a professional health worker (19). Higher degrees of severity among children with severe febrile illness, defined by the symptoms altered consciousness or coma, convulsions, and difficulties in breathing, have also been positively associated with hospital attendance in rural Tanzania (20).

In our study, caregivers mentioned taking children to a PHC for the provider’s professionalism and experience. This may be due to the higher illness severity of these children even though this was not reflected in caregiver’s reported perception of illness severity. However, this data was quantitatively collected and did not explore multidimensional aspects of perceived severity (eg. emotions, knowledge about severe malaria, economic consequences in seeking or not seeking treatment) (21).

Home treatment was provided slightly more often to children brought to a PHC compared to those brought to a CORP (AOR: 1.5). This treatment consisted in most cases of an antipyretic drug which, on its own, would be counter-productive in the treatment of an episode of severe malaria as it would alleviate symptoms and suggest an improvement of the child’s condition. While previous qualitative evidence from Nigeria suggested that children with suspected malaria were usually given paracetamol and an antimalarial or traditional medicine at home (22), home treatment with an antimalarial was uncommon in children in this study.

Children attending PHCs were also more likely to have attended another provider, mostly a drug shop/chemist/pharmacy. While such providers may facilitate access to medicines in a similar way as CORPs, attending such a provider may also unnecessarily delay a visit to a health facility. This is particularly problematic when children suffer from an episode of severe malaria (or another severe illness) and the treatment provided at the drug outlet is inadequate.

Our study found that approximately two-thirds of children with suspected severe malaria did not attend one of the community-based providers on the day or the day after the onset of symptoms. This may in part explain the overall high CFR seen among children in this study. We did not find a significant difference in treatment seeking delay between patients attending CORPs and PHCs even though the latter were more likely to attend another provider beforehand. On the other hand, during COVID-19 pandemic restrictions on movement imposed by the government, delay in seeking treatment increased substantially among those going to a CORP but only marginally among those attending a PHC. CORPs experienced stockouts of commodities and delay in replenishment during the COVID-19 lockdown (data not shown), which could have contributed to discouraging attendance. Considering the retrospective assessment of treatment seeking in this study, the recall period of 28 days may have limited how precisely the time of illness onset and treatment initiation have been remembered. Smaller changes in treatment delay may hence have remained undetected.

Longer delay in attending the first community-based provider was associated with the administration of home treatments. In consideration of the infrequent use of antimalarial medicines at home, any delay in seeking treatment for the episode of suspected severe malaria is therefore likely to have aggravated the severity of the condition. While our study explored treatment seeking delay in number of days, we lack background information on the actual travel time it took and the circumstances of reaching a provider. This may be more relevant in the case of PHCs, which were located outside the patient’s village in 50% of the cases (as opposed to 6% for CORPs). A strenuous transport may negatively affect the condition of a severely sick child.

Other characteristics may have influenced the choice of first community-based provider without necessarily affecting the observed difference in CFR. In Nigeria, PHCs are not located in the most remote areas, although they are often at a considerable distance from the nearest referral health facility (23). This is reflected in our data where children attending a PHC more often resided in urban areas compared to those attending a CORP. Similarly, Druetz *et al*. (2015) showed that in rural areas of Burkina Faso, the decision to consult a community health worker for malaria increased with increased distance to the nearest health facility (24). Provider operating hours also affect health-seeking behaviours. PHCs likely have more standard operating hours and certainly do not provide 24-hour service, while CORPs, who are volunteers and may have other jobs or household activities, may have more flexible hours. We found that on weekends, patients rather attended CORPs than PHCs. A different finding was published by Koce *et al*. (2020) who qualitatively investigated factors influencing self-referral to higher level facilities in Nigeria and demonstrated that the limited staff and operating hours of community-based providers (compared to referral facilities) resulted in patients directly seeking care from higher level facilities (25). Other factors predicting community level provider choice including knowledge of malaria and costs associated with treatment seeking were not assessed.

Taken together, the findings from this analysis suggest that, compared to those attending a CORP, children who first attended a PHC may have been more severely ill, potentially aggravated by inadequate home management, delay in attending the health facility, and possibly a longer and more strenuous travel. Considering that all included children had danger signs of severe illness, caregivers may have underestimated the severity of the child’s condition in many cases.

Together, these factors may have contributed to a higher CFR among children attending a PHC. Other factors affecting the health outcome of children in this study were investigated in previous analyses. For example, the likelihood of death was higher among children who were treated with pre-referral RAS (AOR: 2.42, 95%CI: 1.25 – 4.70), whereas a higher proportion of children first attending PHCs received RAS compared to those attending CORPs (11). This finding may be related to subsequent referral and treatment patterns rather than to RAS itself. For example, only 48% of children completed referral to a Cottage Hospital after administration of RAS by a CORP or PHC (though those attending a PHC were much more likely to complete referral) (12). Post-referral treatment with an injectable antimalarial was common but few children were subsequently treated with the required full course of oral artemisinin-based combination therapy (Signorell et al. manuscript in preparation).

The factors explored in this article in light of other findings are likely to contribute to the high (11.7%) CFR observed in Nigeria but they do not explain the differences between children attending CORPs and those attending PHCs. The higher CFR in those attending a PHC was unexpected since PHCs have higher treatment capacities compared to CORPs.

Previous investigations found an increased risk of dying in the period after the roll-out of RAS in the study area (May 2019 to July 2020) (11), which coincided with the period in which more patients first attended a PHC. This finding is unlikely to be related to RAS itself but may have been influenced by circumstantial changes including the emergence of other infectious diseases with similar clinical presentations. Several suspected viral outbreaks occurred in Adamawa State during the study period, including Lassa Fever and Yellow Fever (26, 27). The symptoms of these diseases are often unspecific and overlap with severe malaria. Differential diagnosis of these diseases was not available for the study patients.

This study provides insight into differences in background characteristics and care seeking between patients attending different community-based providers in Nigeria, contributing to an explanation of the difference in CFRs between patients attending different community-based health care providers in Nigeria. Additionally, it contributes to the limited literature on care seeking for severely ill children, a population which is comparably small and for which published evidence is scarce (21). While findings from this study may not necessarily be directly applicable to other settings, they point to issues that warrant careful attention when promoting the management of severe childhood illness at community level in rural African settings.

## Conclusions

This research provides context on treatment seeking for children with suspected severe malaria between community-based health providers in Nigeria to explain the higher CFRs seen in children visiting PHCs compared to those visiting CORPs in the CARAMAL project. The higher illness severity seen in children visiting PHCs as their first community-based provider likely drives the high CFR.

## Data Availability

The datasets generated during and/or analysed during the current study are available in the Zenodo repository, (DOI: 10.5281/zenodo.5733006), upon reasonable request to the corresponding author.

## List of Abbreviations

ACT: Artemisinin-based Combination Therapy
AOR: Adjusted Odds Ratio
CARAMAL: Community Access to Rectal Artesunate for Malaria
CFR: Case Fatality Rate
CNS: Central Nervous System
CORP: Community Oriented Resource Person
iCCM: integrated Community Case Management
IMCI: Integrated Management of Childhood Illnesses
LGA: Local Government Areas
mRDT: malaria Rapid Diagnostic Test
PHC: Primary Health Center
RAS: Rectal Artesunate
WHO: World Health Organization

## Acknowledgements

We would like to acknowledge the children and their caregivers who participated in the study and the large number of community health providers, data collectors, local coordinators, local CHAI, UNICEF, and WHO colleagues who made this work possible. We additionally would like to thank Amanda Ross for her statistical support.

## Declarations

## Ethics approval and consent to participate

The CARAMAL study protocol was approved by the Research Ethics Review Committee of the World Health Organization (WHO ERC, No. ERC.0003008), the Health Research Ethics Committee of the Adamawa State Ministry of Health (S/MoH/1131/I), the National Health Research Ethics Committee of Nigeria (NHREC/01/01/2007-05/05/2018), and the Scientific and Ethical Review Committee of CHAI (No. 112, 21 Nov 2017). The study is registered on ClinicalTrials.gov (NCT03568344). Provisional verbal informed consent was obtained from caregivers by the community-based provider, while full written informed consent was obtained during a follow-up contact at a referral facility or on day 28. Only patients whose caregivers provided written informed consent were officially enrolled in the study.

## Consent for publication

Not applicable. Results not linkable to individual participants.

## References

1. Kendjo E, Agbenyega T, Bojang K, Newton CR, Bouyou-Akotet M, Pedross F, et al. Mortality patterns and site heterogeneity of severe malaria in African children. PLoS One. 2013;8(3):e58686.

2. Bassat Q, Guinovart C, Sigaúque B, Aide P, Sacarlal J, Nhampossa T, et al. Malaria in rural Mozambique. Part II: children admitted to hospital. Malar J. 2008;7:37.

3. Orimadegun AE, Fawole O, Okereke JO, Akinbami FO, Sodeinde O. Increasing burden of childhood severe malaria in a Nigerian tertiary hospital: implication for control. J Trop Pediatr. 2007;53(3):185–9.

4. World Health Organization. Guidelines for the treatment of malaria – 3rd edition. 2015.

5. Consortium HEFD. Integrated Community Case Management (iCCM) in Sub-Saharan Africa: Successes & Challenges with Access, Speed & Quality. September 2018. Health Economic Finance Development Consortium; 2018.

6. Strides Shasun’s rectal artesunate product receives WHO prequalification. https://www.mmv.org/newsroom/news/strides-shasun-s-rectal-artesunate-product-receives-who-prequalification. Accessed August 23, 2021.

7. Gomes MF, Faiz MA, Gyapong JO, Warsame M, Agbenyega T, Babiker A, et al. Prereferral rectal artesunate to prevent death and disability in severe malaria: a placebo-controlled trial. Lancet. 2009;373(9663):557–66.

8. World Health Organization. World malaria report 2020: 20 years of global progress and challenges. Geneva: World Health Organization; 2020. Report No.: Licence: CC BY-NC-SA 3.0 IGO.

9. Welcome MO. The Nigerian health care system: Need for integrating adequate medical intelligence and surveillance systems. J Pharm Bioallied Sci. 2011;3(4):470–8.

10. Federal Ministry of Health. National Guidelines for Diagnosis and Treatment of Malaria (Third Edition). Abuja; 2015.

11. Hetzel MW, Okitawutshu J, Tshefu A, Omoluabi E, Awor P, Signorell A, et al. Effectiveness of rectal artesunate as pre-referral treatment for severe malaria in children <5 years of age. Preprint at https://doi.org/10.1101/2021.09.24.21263966 (2021).

12. Brunner NC, Omoluabi E, Awor P, Okitawutshu J, Tshefu A, Signorell A, et al. Pre-referral rectal artesunate and referral completion among children with suspected severe malaria in the Democratic Republic of the Congo, Nigeria and Uganda. Preprint at https://doi.org/10.1101/2021.09.27.21264073 (2021).

13. WorldPop. World Population Demographics: Subnational Age/Sex Structures - 2000-2020. https://www.portal.worldpop.org/demographics/. Accessed July 6, 2021.

14. Weber EM, Seaman VY, Stewart RN, Bird TJ, Tatem AJ, McKee JJ, et al. Census-independent population mapping in northern Nigeria. Remote Sens Environ. 2018;204:786–98.

15. Simba DO, Warsame M, Kimbute O, Kakoko D, Petzold M, Tomson G, et al. Factors influencing adherence to referral advice following pre-referral treatment with artesunate suppositories in children in rural Tanzania. Trop Med Int Health. 2009;14(7):775–83.

16. Siribié M, Ajayi IO, Nsungwa-Sabiiti J, Sanou AK, Jegede AS, Afonne C, et al. Compliance With Referral Advice After Treatment With Prereferral Rectal Artesunate: A Study in 3 Sub-Saharan African Countries. Clin Infect Dis. 2016;63](Suppl 5):S283–s9.

17. Ajayi IO, Nsungwa-Sabiiti J, Siribié M, Falade CO, Sermé L, Balyeku A, et al. Feasibility of Malaria Diagnosis and Management in Burkina Faso, Nigeria, and Uganda: A Community-Based Observational Study. Clin Infect Dis. 2016;63](Suppl 5):S245–s55.

18. Warsame M, Gyapong M, Mpeka B, Rodrigues A, Singlovic J, Babiker A, et al. Pre-referral Rectal Artesunate Treatment by Community-Based Treatment Providers in Ghana, Guinea-Bissau, Tanzania, and Uganda (Study 18): A Cluster-Randomized Trial. Clin Infect Dis. 2016;63](Suppl 5):S312–s21.

19. Sauerborn R, Nougtara A, Diesfeld HJ. Low utilization of community health workers: results from a household interview survey in Burkina Faso. Soc Sci Med. 1989;29(10):1163–74.

20. Castellani J, Mihaylova B, Evers SM, Paulus AT, Mrango ZE, Kimbute O, et al. Out-of-pocket costs and other determinants of access to healthcare for children with febrile illnesses: a case-control study in rural Tanzania. PLoS One. 2015;10(4):e0122386.

21. Brunner N, Awor P, Hetzel M. Definitions of Severity in Treatment Seeking Studies of Febrile Illness in Children in Low and Middle Income Countries: A Scoping Review. International Journal of Public Health. 2021;66](74).

22. Elimian KO, Myles PR, Phalkey R, Sadoh A, Pritchard C. ‘Everybody in Nigeria is a doctor…’: a qualitative study of stakeholder perspectives on lay diagnosis of malaria and pneumonia in Nigeria. J Public Health (Oxf). 2020;42(2):353–61.

23. Kuupiel D, Adu KM, Bawontuo V, Mashamba-Thompson TP. Geographical Accessibility to District Hospitals/Medical Laboratories for Comprehensive Antenatal Point-of-Care Diagnostic Services in the Upper East Region, Ghana. EClinicalMedicine. 2019;13:74–80.

24. Druetz T, Ridde V, Kouanda S, Ly A, Diabaté S, Haddad S. Utilization of community health workers for malaria treatment: results from a three-year panel study in the districts of Kaya and Zorgho, Burkina Faso. Malar J. 2015;14:71.

25. Koce FG, Randhawa G, Ochieng B. A qualitative study of health care providers’ perceptions and experiences of patients bypassing primary healthcare facilities: a focus from Nigeria. Journal of Global Health Reports. 2020.

26. World Health Organization. Yellow Fever – Nigeria. Accessed October 19, 2021.

27. Agbonlahor DE, Akpede GO, Happi CT, Tomori O. 52 Years of Lassa Fever Outbreaks in Nigeria, 1969-2020: An Epidemiologic Analysis of the Temporal and Spatial Trends. Am J Trop Med Hyg. 2021.

