## Supplemental materials for "Treatment-seeking for children with suspected severe malaria attending community health workers and primary health centres in Adamawa State, Nigeria"

### Statistical methods and sensitivity analysis

Our research aimed to compare differences between children enrolled by CORPs and PHCs. To obtain correct inferences, we needed to account for the variation in the patients’ characteristics between clusters, that is the enrolling CORPs and PHCs. Standard methods for cluster adjustment such as mixed models and generalized estimating equations (GEEs) with exchangeable correlation structure fail here, because cluster membership determines the outcome. In the main text, we present results using clustered standard errors, an approach re-examined recently in econometrics literature and recommended in settings where the exposure is clustered (1), as is our case. In this appendix, we provide also the estimates from a GEE model with an independent working correlation structure. Note that the GEEs estimate population-averaged (marginal) effect, while logistic regressions the subject-specific (conditional) effect. We find that both methods give results that are broadly in agreement, supporting the conclusions presented in the main text.

Table S1. Estimates generated through multiple models

|  | **Clustered SE** | | |  | **GEE with independent working correlation structure** | | |
| --- | --- | --- | --- | --- | --- | --- | --- |
| **Danger sign** | **AOR*** | **95% CI*** | **p-value*** |  | **AOR*** | **95% CI*** | **p-value*** |
| Convulsions | 3.1 | (2.0 - 4.9) | <0.01 |  | 1.9 | (1.4 - 2.7) | <0.01 |
| Not able to drink or feed anything | 0.8 | (0.6 - 1.2) | 0.28 |  | 0.9 | (0.8 - 1.1) | 0.39 |
| Vomits everything | 0.5 | (0.4 - 0.8) | <0.01 |  | 0.8 | (0.6 - 0.9) | 0.01 |
| Unusually sleepy or unconscious | 2.0 | (1.4 - 2.9) | <0.01 |  | 1.5 | (1.1 - 1.9) | <0.01 |

#### References

1.	Abadie A, Athey S, Imbens GW, Wooldridge J. When should you adjust standard errors for clustering? : National Bureau of Economic Research; 2017.

### Inclusion criteria

Figure S1. Inclusion flow chart

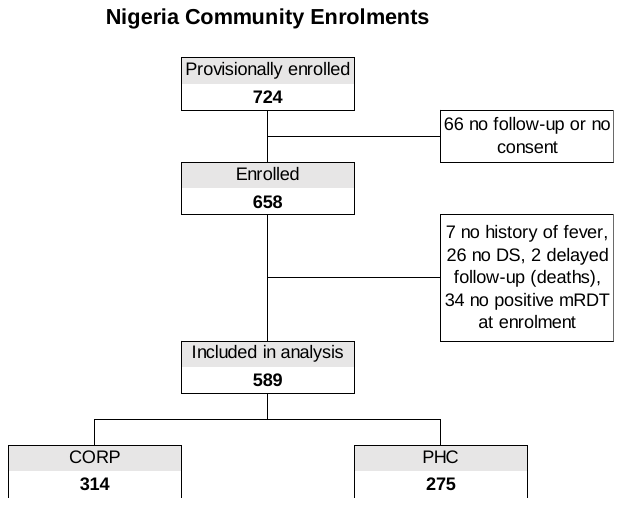

### Unadjusted estimates

Table S2. Study population characteristics by enrolling provider with unadjusted estimates

|  | **CORP** | |  | **PHC** | |  |  |  |  |
| --- | --- | --- | --- | --- | --- | --- | --- | --- | --- |
|  | **N** | **%** |  | **N** | **%** |  | **OR*** | **95% CI*** | **p-value*** |
| **Total** | **314** |  |  | **275** |  |  |  |  |  |
| **Child sex** |  |  |  |  |  |  |  |  |  |
| Male | 188 | 60 |  | 164 | 60 |  | Ref. |  |  |
| Female | 126 | 40 |  | 111 | 40 |  | 1.0 | (0.7 - 1.4) | 0.95 |
| **Child age (years)** |  |  |  |  |  |  |  |  |  |
| 0 | 40 | 13 |  | 27 | 10 |  | Ref. |  | 0.76 |
| 1 | 86 | 27 |  | 74 | 27 |  | 1.3 | (0.6 - 2.5) |  |
| 2 | 82 | 26 |  | 82 | 30 |  | 1.5 | (0.8 - 2.8) |  |
| 3 | 68 | 22 |  | 55 | 20 |  | 1.2 | (0.6 - 2.3) |  |
| 4 | 38 | 12 |  | 37 | 13 |  | 1.4 | (0.6 - 3.2) |  |
| **Caregiver sex** |  |  |  |  |  |  |  |  |  |
| Male | 102 | 32 |  | 95 | 35 |  | Ref. |  |  |
| Female | 212 | 68 |  | 180 | 65 |  | 0.9 | (0.6 - 1.3) | 0.64 |
| **Caregiver age** |  |  |  |  |  |  |  |  |  |
| < 25 | 42 | 13 |  | 46 | 17 |  | Ref. |  | 0.47 |
| 25 - 34 | 167 | 53 |  | 128 | 47 |  | 0.7 | (0.4 - 1.2) |  |
| 35 - 45 | 75 | 24 |  | 72 | 26 |  | 0.9 | (0.5 - 1.6) |  |
| ≥ 45 | 30 | 10 |  | 28 | 10 |  | 0.9 | (0.4 - 1.6) |  |
| Missing | 0 | 0 |  | 1 | 0 |  | - |  |  |
| **Caregiver education**** |  |  |  |  |  |  |  |  |  |
| Never attended school | 54 | 17 |  | 65 | 24 |  | Ref. |  | 0.63 |
| Primary or lower | 15 | 5 |  | 26 | 9 |  | 1.4 | (0.6 - 3.3) |  |
| Secondary or higher | 28 | 9 |  | 44 | 16 |  | 1.3 | (0.6 - 2.6) |  |
| Quranic | 22 | 7 |  | 39 | 14 |  | 1.5 | (0.7 - 3.1) |  |
| Missing | 195 | 62 |  | 101 | 37 |  | - |  |  |
| **LGA** |  |  |  |  |  |  |  |  |  |
| Fufore | 143 | 46 |  | 94 | 34 |  | Ref. |  | 0.12 |
| Mayo-Belwa | 95 | 30 |  | 154 | 56 |  | 2.5 | (0.6 - 11.0) |  |
| Song | 76 | 24 |  | 27 | 10 |  | 0.5 | (0.1 - 2.0) |  |
| **Residence** |  |  |  |  |  |  |  |  |  |
| Rural | 298 | 95 |  | 227 | 83 |  | Ref. |  |  |
| Urban | 14 | 4 |  | 43 | 16 |  | 4.0 | (1.2 - 13.5) | 0.02 |
| Missing | 2 | 1 |  | 5 | 2 |  | - |  |  |
| **Day of enrolment** |  |  |  |  |  |  |  |  |  |
| Workday | 241 | 77 |  | 239 | 87 |  | Ref. |  |  |
| Weekend | 73 | 23 |  | 36 | 13 |  | 0.5 | (0.3 - 0.9) | 0.02 |
| **Season** |  |  |  |  |  |  |  |  |  |
| Dry season | 83 | 26 |  | 66 | 24 |  | Ref. |  |  |
| Rainy season | 231 | 74 |  | 209 | 76 |  | 1.1 | (0.7 - 1.9) | 0.63 |
| **RAS implementation phase** |  |  |  |  |  |  |  |  |  |
| Pre-RAS | 156 | 50 |  | 61 | 22 |  | Ref. |  |  |
| Post-RAS | 158 | 50 |  | 214 | 78 |  | 3.5 | (1.8 - 6.7) | <0.01 |
| **COVID-19 pandemic** |  |  |  |  |  |  |  |  |  |
| Pre-COVID-19 | 268 | 85 |  | 207 | 75 |  | Ref. |  |  |
| COVID-19 period | 46 | 15 |  | 68 | 25 |  | 1.9 | (1.0 - 3.6) | 0.04 |

*Unadjusted logistic regression with standard errors clustered at the level of the health care provider. Likelihood ratio test used to calculate p-values for categorical variables. **Data not collect in collected during the complete study period.

Table S3. Signs and symptoms of disease and caregiver’s perceived severity of the illness with unadjusted estimates

|  | **CORP** | |  | **PHC** | |  |  |  |  |
| --- | --- | --- | --- | --- | --- | --- | --- | --- | --- |
|  | **N** | **%** |  | **N** | **%** |  | **OR*** | **95% CI*** | **p-value*** |
| **Total** | **314** |  |  | **275** |  |  |  |  |  |
| **RAS danger sign** |  |  |  |  |  |  |  |  |  |
| Convulsions | 170 | 54 |  | 217 | 79 |  | 3.2 | (2.0 - 5.0) | <0.01 |
| Not able to drink or feed anything | 200 | 64 |  | 159 | 58 |  | 0.8 | (0.5 - 1.1) | 0.18 |
| Vomits everything | 227 | 72 |  | 160 | 58 |  | 0.5 | (0.4 - 0.8) | <0.01 |
| Unusually sleepy or unconscious | 175 | 56 |  | 193 | 70 |  | 1.9 | (1.3 - 2.7) | <0.01 |
|  | **117** |  |  | **172** |  |  |  |  |  |
| Yellowness of the eyes ** | 8 | 7 |  | 4 | 2 |  | 0.3 | (0.1 - 1.1) | 0.06 |
| **Other danger signs** | **314** |  |  | **275** |  |  |  |  |  |
| Blood in stool | 57 | 18 |  | 20 | 7 |  | 0.4 | (0.2 - 0.6) | <0.01 |
| Swelling of both feet | 5 | 2 |  | 11 | 4 |  | 2.6 | (0.8 - 8.6) | 0.12 |
| Unable to sit or stand | 140 | 45 |  | 147 | 53 |  | 1.4 | (1.0 - 2.0) | 0.04 |
|  | **117** |  |  | **172** |  |  |  |  |  |
| Cough for 14 days or more ** | 3 | 3 |  | 3 | 2 |  | 0.7 | (0.1 - 3.2) | 0.62 |
| Diarrhoea for 14 days or more ** | 4 | 3 |  | 1 | 1 |  | 0.2 | (0.0 - 1.3) | 0.09 |
| Fever lasting 7 days or more ** | 57 | 49 |  | 53 | 31 |  | 0.6 | (0.4 - 1.0) | 0.05 |
| Whiteness of the palms and sole ** | 3 | 3 |  | 14 | 8 |  | 3.4 | (0.9 - 12.6) | 0.07 |
| Coke coloured urine** | 26 | 22 |  | 48 | 28 |  | 1.4 | (0.7 - 2.7) | 0.38 |
| Number of danger signs *** | **314** |  |  | **275** |  |  |  |  |  |
| 0 - 1 | 52 | 17 |  | 32 | 12 |  | Ref. |  | 0.06 |
| 2 - 3 | 138 | 44 |  | 106 | 39 |  | 1.2 | (0.7 - 2.1) |  |
| ≥ 4 | 124 | 39 |  | 137 | 50 |  | 1.8 | (1.1 - 3.0) |  |
| **CNS  danger sign (unusually sleepy or unconscious or convulsions)** | 231 | 74 |  | 248 | 90 |  | 3.3 | (1.9 - 5.7) | <0.01 |
| **Perceived severity of the illness** |  |  |  |  |  |  |  |  |  |
| Not fatal | 236 | 75 |  | 193 | 70 |  | Ref. |  |  |
| Fatal | 76 | 24 |  | 79 | 29 |  | 1.3 | (0.8 - 2.0) | 0.28 |
| Don’t know/Missing | 2 | 1 |  | 3 | 1 |  | - |  |  |

*Unadjusted logistic regression with standard errors clustered at the level of the health care provider. Likelihood ratio test used to calculate p-values for categorical variables. **3 children were not said to have any danger signs by caregivers at follow-up but health providers noted danger signs at enrolment. ***Only includes danger signs collected across the whole study period.

Table S4. Actions taken at home prior to consulting health providers

|  | **CORP** | |  | **PHC** | |  |  |  |  |
| --- | --- | --- | --- | --- | --- | --- | --- | --- | --- |
|  | **N** | **%** |  | **N** | **%** |  | **OR*** | **95% CI*** | **p-value*** |
| **Total** | **314** |  |  | **275** |  |  |  |  |  |
| **Any home treatment** |  |  |  |  |  |  |  |  |  |
| No | 209 | 67 |  | 157 | 57 |  | Ref. |  |  |
| Yes | 103 | 33 |  | 116 | 42 |  | 1.5 | (1.0 - 2.2) | 0.03 |
| Missing | 2 | 1 |  | 2 | 1 |  | - |  |  |
| **Actions taken** | **103** |  |  | **116** |  |  |  |  |  |
| Traditional medicines/herbs | 22 | 21 |  | 26 | 22 |  | 1.1 | (0.5 - 2.2) | 0.87 |
| Tepid sponging | 7 | 7 |  | 17 | 15 |  | 2.4 | (0.9 - 6.4) | 0.09 |
| Given medicine | 79 | 77 |  | 85 | 73 |  | 0.8 | (0.5 - 1.5) | 0.55 |
| **Caregiver remembers what medicine the child received** | **79** |  |  | **85** |  |  |  |  |  |
| No | 12 | 15 |  | 9 | 11 |  | Ref. |  |  |
| Yes | 67 | 85 |  | 76 | 89 |  | 1.5 | (0.6 - 4.0) | 0.40 |
| **Medicines given** | **67** |  |  | **76** |  |  |  |  |  |
| Artemether-lumefantrine | 10 | 15 |  | 11 | 14 |  | 1.0 | (0.3 - 3.1) | 0.95 |
| Artesunate-amodiaquine | 0 | 0 |  | 0 | 0 |  | - |  | - |
| Oral rehydration solution (ORS) | 1 | 1 |  | 1 | 1 |  | 0.9 | (0.1 - 12.9) | 0.93 |
| Paracetamol | 57 | 85 |  | 70 | 92 |  | 2.0 | (0.6 - 6.9) | 0.25 |
| Other | 6 | 9 |  | 5 | 7 |  | 0.7 | (0.2 - 2.6) | 0.61 |

*Unadjusted logistic regression with standard errors clustered at the level of the health care provider. Likelihood ratio test used to calculate p-values for categorical variables.
